# Rapid-Response Viral Genome Detection using TWIST Capture and Nanopore Flongle Sequencing

**DOI:** 10.64898/2026.06.18.26355521

**Authors:** Annabel Rector, Mandy Bloemen, Jill Swinnen, Mustafa Karataş, Lander De Coninck, Jelle Matthijnssens, Marc Van Ranst, Elke Wollants

## Abstract

**Background:** Rapid detection of viral pathogens can be challenging, especially when routine PCR fails. Conventional assays typically detect known viruses which are specifically targeted by the assay, which may result in the failure to identify novel or non-targeted viruses. Broad-range hybrid-capture sequencing enables unbiased detection of viruses, including those that are uncommon or divergent.

**Methods:** We combined the TWIST Comprehensive Viral Research Panel (>3,000 virus species) with Oxford Nanopore Flongle sequencing for easy and quick viral genome detection. The workflow includes random-primed cDNA synthesis, dsDNA conversion, TWIST probe enrichment, and Nanopore sequencing. Performance was evaluated using the QCMD 2024 Viral Metagenomics EQA panel and one clinical sample.

**Results:** All expected targets of the QCMD 2024 Viral Metagenomics EQA panel were detected; eight of thirteen viruses achieved ≥90% genome coverage. The negative control showed no targeted viral reads. Mixed infections of DNA and RNA viruses were resolved accurately. The workflow from nucleic acid extraction to obtaining sequence data was completed within 3 days.

**Conclusions:** Combination of Flongle and hybrid-capture offers a rapid, sensitive sequencing method for detecting diverse viral genomes, suitable for outbreak response, urgent diagnostics, and surveillance of multiple divergent viruses at once.

## Introduction

Detecting unexpected or low-abundance viral agents in clinical and environmental specimens remains a diagnostic challenge when routine molecular tests yield negative results despite strong suspicion of infection. Common diagnostic assays such as quantitative real-time PCR (qRT-PCR) target specific pathogens, relying on a priori knowledge of the targeted nucleotide sequence, and therefore cannot reveal novel, divergent viral variants or species. These assays often focus on detection of a single virus or a limited number of viruses which are most commonly associated with the syndrome under investigation, which may result in less apparent or rarer causes being overlooked. An alternative could be untargeted metagenomic sequencing, which in theory allows for unbiased and hypothesis-free detection of an infectious agent. Its application is however hindered by the presence of high levels of contaminating human host and/or (commensalic) bacterial nucleic acids in patient samples, resulting in reduced analytical sensitivity (1). Broad, probe-based hybrid-capture sequencing offers an attractive complementary strategy, simultaneously enriching thousands of viral taxa and enabling unbiased genome recovery while maintaining a high sensitivity (2).

The TWIST Comprehensive Viral Research Panel (CVRP) contains 1,052,421 unique probes targeting 3,153 human-infective virus species, allowing comprehensive capture of both known and emerging viral agents. The hybridisation-based target enrichment enables complete characterization of known as well as novel viral sequences, in combination with a high sensitivity (3). The TWIST CVRP has previously been evaluated in combination with short-read sequencing such as Illumina and Aviti (2,4,5). We wanted to explore the possibility of coupling TWIST CVRP to a rapid sequencing platform such as the Oxford Nanopore Technologies (ONT) Flongle, allowing same-day viral genome detection after hybrid capture from one or a few samples. The small-format Flongle flow cell is particularly suitable for urgent “rescue” analyses in which results are required quickly but the number of specimens is too limited for high-throughput sequencing runs. Here we describe a streamlined hybrid-capture and sequencing workflow that employs the TWIST CVRP for enrichment, followed by Nanopore ligation sequencing on a Flongle flow cell. It covers all viral genome types (ssRNA, dsRNA, ssDNA, dsDNA) through random-primed cDNA synthesis and double-stranded DNA conversion, followed by TWIST enrichment and ONT sequencing.

To evaluate the performance and analytical sensitivity of the viral metagenomics workflow, external quality assessment (EQA) samples were obtained from the *Quality Control for Molecular Diagnostics (QCMD)* program (www.qcmd.org). The QCMD panels consisted of well-characterized positive and negative specimens containing known viral targets in a clinically relevant matrix. The suitability for clinical practice was assessed by testing a patient sample using the TWIST panel in combination with Illumina sequencing and ONT Flongle sequencing in parallel.

## Materials and Methods

### Samples

QCMD samples were extracted with the RNeasy Mini Kit (Cat. No. 74106, Qiagen). A respiratory sample obtained from an infant who died of sudden infant death syndrome (SIDS) was extracted as previously described (6).

### cDNA synthesis and second-strand synthesis: ∼3 hours

Since the TWIST protocol is compatible with ssRNA, dsRNA, ssDNA, and dsDNA, an initial cDNA synthesis step is required. This is achieved via sequential first- and second-strand synthesis. To generate the first cDNA strand, 5 µL of random hexamer primers (Cat. No. S1230S, New England Biolabs) was added to 15 µL nucleic acid extract, followed by incubation at 95°C for 5 minutes. Subsequently, 25 µL of ProtoScript II Reaction Mix and 5 µL of ProtoScript II Enzyme Mix (Cat. No. 6560L, New England Biolabs) were added to each reaction, yielding a final volume of 50 µL. The reaction was incubated for 5 minutes at 25°C, 60 minutes at 42°C, and 5 minutes at 80°C.

Second-strand synthesis was performed by adding a mixture consisting of 18 µL nuclease-free water, 8 µL NEBNext Second Strand Synthesis Reaction Buffer, and 4 µL NEBNext Second Strand Synthesis Enzyme Mix (Cat. No. E6111, New England Biolabs) to 50 µL of the completed first-strand reaction, resulting in a reaction volume of 80 µL. This was incubated at 16°C for 60 minutes. The synthesized cDNA was purified using DNA purification beads (Cat. No. 100573, TWIST BioScience) at a 1.2x bead-to-sample ratio and quantified with the Qubit™ 1X dsDNA HS Assay Kit (Cat. No. Q33231) using a Qubit fluorometer.

### Library preparation: ∼3 hours

Enzymatic fragmentation, end-repair, and dA-tailing were performed in preparation for adapter ligation, using reagents from the TWIST Library Preparation EF Kit (Cat. No. 104206, TWIST BioScience). For each reaction, 25 µL of cDNA (≥50 ng) was combined with a fragmentation mix consisting of 15 µL nuclease-free water, 4 µL Frag/AT Buffer, and 6 µL Frag/AT Enzymes. The reaction was incubated for 5 minutes at 30°C, followed by 30 minutes at 65°C.

Subsequently, 2.5 µL of TWIST Universal Adapters (Cat. No. 101307, TWIST BioScience) was added to the dA-tailed DNA fragments. A ligation mix containing 2.5 µL of nuclease-free water and 20 µL Ligation Master Mix was then added to each sample. The ligation reaction was incubated at 20°C for 15 minutes and purified using DNA purification beads at a 0.8x bead-to-sample ratio.

To complete library preparation, adapter-ligated libraries were amplified using TWIST UDI primers (Cat. No. 101307, TWIST BioScience). For each reaction, 10 µL primers and 25 µL of Equinox Library Amp Mix (2x) (Cat. No. 104178, TWIST BioScience) were added to the purified cDNA libraries. The amplification conditions were: initial denaturation at 98°C for 45 seconds, followed by 17 cycles of 98°C for 15 seconds, 60°C for 30 seconds, and 72°C for 30 seconds, with a final extension at 72°C for 1 minute. Libraries were purified using a 1x bead ratio and quantified with the Qubit™ 1X dsDNA HS Assay Kit (Cat. No. Q33231) using a Qubit fluorometer.

### Target enrichment and amplification: ∼20 hours

For target enrichment, indexed libraries from the previous step were pooled to obtain 400 ng per library. The pooled DNA was concentrated using DNA purification beads at a 1.8x ratio and eluted in a solution containing 7 µL Universal Blockers and 5 µL Blocker Solution (Cat. No. 100578, TWIST BioScience).

Hybridization was performed using reagents from the TWIST Hybridization and Wash Kit (Cat. No. 104178, TWIST BioScience). The hybridization mix was pre-heated at 65°C for 10 minutes and cooled to room temperature for 5 minutes. The probe mix was prepared by combining 20 µL Hybridization Mix, 4 µL TWIST Fixed Panel, and 4 µL nuclease-free water, then incubated at 95°C for 2 minutes and immediately cooled to 4°C for 5 minutes. During this cooling step, the pooled indexed libraries were also heated at 95°C for 5 minutes. Both probe and library mixes were equilibrated to room temperature for 5 minutes before combining 28 µL of probe solution with 12 µL of library mix giving it a total reaction volume of 40 µL. Subsequently, 30 µL Hybridization Enhancer was added, and hybridization was carried out at 70°C for 17 hours with the thermocycler lid heated to 85°C.

Following hybridization, targets were captured using streptavidin beads. Prior to binding, Binding Buffer (BB) (Cat. No. 104325, TWIST BioScience), Wash Buffer 1 (WB1), and Wash Buffer 2 (WB2) (Cat. No. 104178, TWIST BioScience) were pre-warmed to 48°C to dissolve any precipitates. BB and WB1 were cooled to room temperature, while WB2 was maintained at 48°C. For each pool, 100 µL of streptavidin beads (Cat. No. 104325, TWIST BioScience) were washed three times using 200 µL BB. After the last washing step, the streptavidin beads were resuspended in 200 µL BB and 70 µL of the hybridized library mix was added. This mixture was incubated at room temperature for 30 minutes using a shaker to keep the solution mixed, followed by magnetic separation and removal of the supernatant. Beads were sequentially washed with WB1 and WB2, and resuspended in 45 µL nuclease-free water.

Post-capture PCR amplification was performed by mixing 22.5 µL of bead-bound DNA with 2.5 µL ILMN amplification primers (Cat. No. 104178, TWIST BioScience) and 25 µL Equinox Library Amp Mix 2x (Cat. No. 104178, TWIST BioScience). Thermocycling conditions were as follows: initial denaturation at 98°C for 45 seconds; 12 cycles of 98°C for 15 seconds, 60°C for 30 seconds, and 72°C for 30 seconds; and a final extension at 72°C for 1 minute. Amplified libraries were purified using a 1x bead ratio, analyzed using the Agilent Bioanalyzer High Sensitivity DNA Kit, and quantified with the Qubit™ 1X dsDNA HS Assay Kit (Cat. No. Q33231) on a Qubit fluorometer.

### ONT ligation library preparation: ∼2 hours

To prepare the library for ligation sequencing, a second DNA repair and end-prep step was performed by adding 100 fmol of enriched DNA to 1.75 µL NEB FFPE DNA Repair Buffer (Cat. No. E6622AA, New England Biolabs) and 0.75 µL Ultra II End-Prep Enzyme Mix (Cat. No. E7646A, New England Biolabs). The mixture was then brought to a total volume of 15 µL with nuclease-free water and incubated for 5 minutes at 20°C, followed by 5 minutes at 65°C. Subsequently, the mix was purified using a 1:1 ratio of AMPure XP beads and the concentration was measured using a Qubit fluorometer.

Following the end-prep, an adaptor ligation was performed on 75 ng of DNA from the previous step using the ONT Ligation Sequencing Kit V14 (SQK-LSK114, Oxford Nanopore Technologies) by adding 2.5 µL Ligation Adapter, 2.5 µL Quick Ligation Buffer (Cat. No. E6056S, New England Biolabs), and 5 µL Quick T4 Ligase (Cat. No. E6056S, New England Biolabs). The ligation was carried out for 20 minutes at 20°C. Afterwards, the reaction was purified using AMPure XP beads, using Short Fragment Buffer (SQK-LSK114, Oxford Nanopore Technologies) as WB. The eluate was quantified on a Qubit fluorometer.

### Flongle sequencing: up to 24 hours

For loading onto an R10.4.1 Flongle flow cell (FLO-FLG114, Oxford Nanopore Technologies), the library was prepared in Elution Buffer (SQK-LSK114) to a total DNA amount of 4 ng. After that, the sequencing mix was made adding 15 µL Sequencing Buffer (Flongle Sequencing Expansion kit EXP-FSE002, Oxford Nanopore Technologies) and 10 µL Library Beads (EXP-FSE002) to 5 µL of diluted library, yielding a total volume of 30 µL. The Flongle flowcell was primed with a mix of 117 µL Flow Cell Flush (EXP-FSE002) and 3 µL Flow Cell Tether (SQK-LSK114) after which the sequencing mix was loaded into the Flongle flow cell according to the ONT protocol.

Sequencing was performed on a Mk1C device (Oxford Nanopore Technologies). Super-accurate basecalling and de-multiplexing of the sequencing reads were performed post-sequencing in MinKNOW version 25.05.14 with basecalling model v5.0.0 - 400 bps. Fastq reads with a minimum Q-score of 10 were used for subsequent analysis.

### Genome assembly and viral classification

Per sample, raw sequencing reads were analyzed using the *Genome Detective Virus Tool* version 2.21.4 (https://www.genomedetective.com) (7). This easy to use platform automatically performs quality control, read trimming, and both de novo and reference-based assembly of viral genomes. Resulting contigs were taxonomically classified using the Genome Detective reference database through a BLAST-based similarity search. Consensus genomes and gene annotations were generated, and summary reports were used to confirm species- and strain-level identification.

## Results

### Detection and sequencing performance of QCMD 2024 EQA samples

All six samples from the *QCMD 2024 Viral Metagenomics External Quality Assessment (EQA)* panel were multiplexed in a single experiment and processed using a Flongle flow cell with 77 active pores at start of sequencing. A total of 2.06 million reads (886.71 Mb called bases) were generated, and analyzed via the *Genome Detective* pipeline. Results are shown in table 1. The metagenomic workflow successfully identified all expected viruses across the 2024 QCMD panel, with genome coverage exceeding 90% for 8 of 13 detected viruses. The negative control (sample 5) showed no targeted viral reads, confirming assay specificity. Mixed infections (e.g., Samples 1, 2, 4, and 6) were all correctly identified, demonstrating robustness of the workflow in multiplexed contexts. These results confirm both the sensitivity and specificity of the approach for detecting diverse DNA and RNA viruses in mixed and low-concentration samples.

**Table 1.** QCMD EQA samples sequencing result.

|  | Nanopore sequencing using Flongle |  |  |  | QCMD report |  |
| --- | --- | --- | --- | --- | --- | --- |
| Sample | Results<br>Genome Detective | #<br>Reads | Mean<br>Depth of<br>Coverage | %<br>Genome<br>Coverage | QCMD 2024<br>Synthetic CSF<br>with human cells | Viral load<br>(Log10<br>copies/ml) |
| NGSmeta<br>24S-01 | Monkeypox virus<br>(West African clade) | 126,419 | 153.4 | 99.9 | Monkeypox | 4 |
|  | HIV-1 Subtype B | 389 | 11.9 | 96.3 | HIV-1 | 4 |
| NGSmeta<br>24S-02 | Herpes simplex virus<br>type 2 | 43,106 | 78.0 | 92.4 | HSV-2 | 3.4 |
|  | Influenza A (H3N2) | 3,545 | 63.7 | 90.0 | Influenza A H3N2 | 5.7 |
|  | Enterovirus D68-B3 | 3,152 | 104.6 | 97.9 | Enterovirus D68 | 4.5 |
| NGSmeta<br>24S-03 | Cytomegalovirus<br>(CMV) | 8,002 | 25.3 | 32.8 | Cytomegalovirus | 4 |
| NGSmeta<br>24S-04 | Enterovirus D68-B3 | 3,793 | 132.6 | 95.4 | Enterovirus D68 | 4.5 |
|  | Herpes simplex virus<br>type 1 | 5,681 | 24.7 | 38.4 | HSV-1 | 3.4 |
|  | Monkeypox virus<br>(West African) | 8,113 | 23.1 | 41.0 | Monkeypox | 3 |
| NGSmeta<br>24S-05 | Negative | — | — | — | negative | N/A |
| **NGSmeta**<br>**24S-06** | Enterovirus D68-B3 | 5,058 | 178.4 | 97.1 | Enterovirus D68 | 4.5 |
|  | Herpes simplex virus<br>type 1* | 799 | 16.8 | 9.1 | HSV-1 | 1.4 |
|  | Herpes simplex virus<br>type 2* | 742 | 17.1 | 7.9 | HSV-2 | 1.4 |

\* These detections were identified under the "discoveries" section of the Genome Detective report rather than definitive detections list, which may mean significantly lower horizontal coverage and abundance compared to other viruses

### Detection and sequencing performance of a patient sample

A patient sample was investigated using the Twist protocol followed by sequencing with Illumina and ONT Flongle in parallel. Using Illumina sequencing, 4 respiratory pathogens were detected in this single sample, as previously described (6). The comparison of the results of Illumina and ONT Flongle are summarized in Table 2. As expected, Illumina sequencing achieved higher genome coverage possibly due to its greater sequencing depth. However, the data originating from the Flongle run provided accurate identification of all 4 pathogens present in the sample.

**Table 2.** SIDS Respiratory sample sequencing result.

| Nanopore sequencing using Flongle |  |  | Illumina sequencing |  |  |
| --- | --- | --- | --- | --- | --- |
| Results<br>Genome Detective | Reads | %<br>Genome<br>Coverage | Illumina<br>Esviritu<br>0.2.3 | Reads | %<br>Genome<br>coverage |
| HHV-4 (Epstein-Barr virus) | 3795 | 92 | EBV | 49643 | 96 |
| Rhinovirus-A12 | 2027 | 96 | HRV-A12 | 10060 | 100 |
| Enterovirus-C105 | 967 | 66 | EV-C105 | 9877 | 100 |
| Parechovirus-1B | 359 | 70 | HPeV-1 | 526 | 37 |

## Discussion

We developed a rapid hybrid-capture Nanopore sequencing method for urgent viral genome detection in previously tested but unresolved samples. The workflow combines the broad coverage of the TWIST CVRP with the speed and agility of ONT Flongle sequencing, allowing identification of thousands of human and animal viruses. This approach can result in a significant time gain compared to Illumina sequencing, where multiple samples typically need to be pooled for cost-effectiveness, and the overall run time is usually considerably longer. ONT sequencing moreover provides real-time access to sequencing data, which allows preliminary analysis of the results even before the run is completed. This could be an advantage for samples with high viral loads, where a sufficient amount of reads to allow virus identification can already be available after short runtimes.

The Flongle flow cell in combination with a small-scale sequencing device provides a sequencing strategy that is accessible for most laboratories, avoiding the need of time-consuming outsourcing to centralized sequencing labs. We performed the analysis on a user-friendly online platform, eliminating the need for developing in-house bioinformatics pipelines. This makes the workflow widely deployable across laboratories. Our low-throughput but sensitive, flexible workflow is ideally suited for time-critical investigations such as outbreak tracing, unexpected clinical cases, or environmental surveillance, where immediate sequencing of a few samples can provide decisive information.

## Data Availability

All data produced in the present study are available upon reasonable request to the authors

## Acknowledgements

We thank UZ Leuven for the QCMD samples for viral metagenomics.

